# Post-marketing active surveillance of Guillan Barré Syndrome following vaccination with anti-COVID-19 vaccines in persons aged ≥12 years in Italy: a multi-database self-controlled case series study

**DOI:** 10.1101/2023.01.17.23284585

**Authors:** Cristina Morciano, Stefania Spila Alegiani, Francesca Menniti Ippoliti, Valeria Belleudi, Gianluca Trifirò, Giovanna Zanoni, Aurora Puccini, Ester Sapigni, Nadia Mores, Olivia Leoni, Giuseppe Monaco, Elena Clagnan, Cristina Zappetti, Emanuela Bovo, Roberto Da Cas, Marco Massari

**Author notes:** Contributed equally to this work as first authors (Cristina Morciano, Stefania Spila Alegiani).

## Abstract

**Background:** Case reports of Guillain Barrè syndrome (GBS) following the Coronavirus Disease 2019 (COVID-19) vaccines administration have been reported. This study investigated the risk of GBS after vaccination with anti-COVID-19 vaccines (BNT162b2/Tozinameran; mRNA-1273/Elasomeran, ChAdOx1-S and Ad26.COV2-S) in the population aged ≥12 years in Italy.

**Methods:** We conducted a self-controlled case series study (SCCS) using national data on COVID-19 vaccination linked to emergency care/hospital discharge databases. The outcome was the first diagnosis of GBS between 27 December 2020 and 30 September 2021.

Exposure risk period were days 0 (vaccination day) through 42 days following each of the 2 vaccine doses. The remaining periods were considered as non at risk (baseline) period.

The SCCS model, adapted to event-dependent exposures, was fitted using unbiased estimating equations to estimate relative incidences (RIs) and excess of cases (EC) per 100,000 vaccinated by dose and vaccine product. Calendar period was included as time-varying confounder in the model.

**Results:** The study included 15,986,009 persons who received at least one dose of Covid-19 vaccine. During the 42-day risk interval there were a total of 67 cases of GBS after the first dose and 41 cases after the second dose. In the 42-day risk interval, increased risks were observed after the administration of first dose (RI=6.83; 95% CI 2.14-21.85) and second dose (RI=7.41; 95% CI 2.35-23.38) for mRNA-1273 vaccine, corresponding to 0.4 and 0.3 EC per 100,000 vaccinated, respectively. Increased risk was also observed after the first dose of ChAdOx1-S vaccine (RI=6.52; 95% CI 2.88-14.77), corresponding to 1.0 EC per 100,000 vaccinated. There was no evidence of increased risk of GBS after vaccination with BNT162b2 and Ad26.COV2-S vaccines.

In the subgroup analysis by sex an increased risk of GBS was observed among both males and females after mRNA-1273 vaccine. in males an increased risk was observed after the first dose, with a borderline significance (RI=5.26; 95% CI 0.94-29.42; p=0.06) and the second dose (RI=16.50; 95% CI 3.01-90.56) and in females after the first dose (RI=13.44; 95% CI 2.83-63.80). There was also evidence of an increased risk after a first dose of ChAdOx1-S in males (RI=4.94; 95% CI 1.84-13.28) and females (RI=7.14; 95% CI 1.94-26.19).

In the subgroup analysis by age, there was evidence of an increased risk of GBS with mRNA-1273 vaccine among those aged ≥60 years after the first (RI=8.03; 95% CI 2.08-31.03) and second dose (RI=7.71; 95% CI 2.38-24.97). After a first dose of ChAdOx1-S there was evidence of an increased risk of GBS in those aged 40-59 (RI=4.50; 95% CI 1.37-14.79) and in those aged ≥60 years (RI=6.84; 95% CI 2.56-18.28). There was no evidence of increased risk of GBS after vaccination with BNT162b2 and Ad26.COV2-S vaccines in the subgroup analysis by age and sex.

Study limitations include that the outcome was not validated through review of clinical records, the possibility of time-dependent residual confounding and the imprecision of the obtained estimates in the subgroup analysis due to the very low number of events.

**Conclusions:** It is important the continuous monitoring of the suspected adverse events of the COVID-19 vaccines as key component of any vaccination program. Results from this large SCCS study showed an increased risk of GBS after first and second dose of mRNA-1273 and first dose of ChAdOx1-S. However, these findings were compatible with a small number of EC. Our data are reassuring regarding BNT162b and Ad26.COV2-S vaccines with respect to GBS outcome. No increased risk of GBS was detected following each of BNT162b vaccine dose nor any increased risk after Ad26.COV2-S vaccine dose.

## Introduction

The Istituto Superiore di Sanità (National Institute of Health) and the Agenzia Italiana del Farmaco (Italian Medicines Agency) coordinate a national active post-marketing surveillance on effectiveness and safety of COVID-19 vaccines since December 2020.^1^

The active surveillance is based on a multi-database (multi-regional) observational cohort and Self-Controlled Case Series study design using *TheShinISS*, an R-based open-source statistical tool, developed by researchers of the Istituto Superiore di Sanità.^2^ Five Italian Regions (Lombardia, Veneto, Friuli Venezia Giulia, Emilia Romagna, Lazio), representing about 44% of the Italian population aged 12 years or older, are included in the study. As part of this active surveillance, we evaluated the risk of Guillain Barré Syndrome (GBS) after vaccination with anti-COVID-19 vaccines (BNT162b2-Tozinameran, mRNA-1273-Elasomeran, ChAdOx1-S and Ad26.COV2-S) in the population aged ≥12 years, during the ongoing Italian anti-Covid-19 vaccination campaign, based on data from 27 December 2020 to 30 September 2021.

## Methods

### Data source

The active surveillance is based on a dynamic multi-regional observational cohort. A distributed analysis framework is applied using TheShinISS, an R-based open-source statistical tool, developed by the researchers of the National Institute of Health,^2^ that locally processes data collected and updated periodically from regional health care databases according to ad hoc, study-tailored, common data model.

Data on vaccination exposure, on hospitalization for GBS and subjects’ characteristics were retrieved from several routinely collected regional healthcare databases:

- <u>COVID-19 vaccination registry</u> to identify information on administered vaccines (product, date of administration and doses for all vaccinated subjects);
- <u>population registry</u> to identify information on age, sex and vital status (causes of death are not recorded in this registry);
- <u>hospital discharge and emergency care visit databases</u> to identify GBS events in the period pre- and post-vaccination, and information on the comorbidities of the study subjects in the period preceding the vaccination;
- <u>pharmacy claims and copayment exemptions databases</u> to obtain information on the comorbidities of the study subjects in the period preceding the vaccination;
- <u>vaccination registry</u> to identify other vaccinations (e.g., flu and pneumococcal vaccines) administered in the period pre- and post-anti COVID-19 vaccination;
- <u>COVID-19 surveillance system</u> to obtain information on SARS-CoV2 infection and related outcomes.

### Study design

We used a Self-Controlled Case Series (SCCS) design.^3-7^ The SCCS design has emerged as a key methodology for studying the safety of vaccines. This approach only requires information from individuals who have experienced the event of interest, and automatically controls for multiplicative time-invariant confounders, even when these are unmeasured or unknown. Originally designed to analyze the association between vaccination and specific events under the key assumption that events do not influence post-event exposures, this method has been adapted to event-dependent exposures, for example when occurrence of an event may preclude any subsequent exposure (SCCS method for censored, perturbed or curtailed post-event exposures).^6-8^ This is the case in observational studies of vaccines when the event of interest could be a contraindication to vaccination.

By using the adapted SCCS method for event-dependent exposures, we estimated the Relative Incidence of GBS following pre-specified windows at risk after vaccination, in a within-person comparison of different time-periods. The method allows for the control of all time-independent characteristics of subjects. The SCCS method allows also for adjustment of potential time-varying confounders such as seasonal variation in risks.

### Study period and population

We investigated the association between anti-COVID-19 vaccines and subsequent onset of GBS in the population aged ≥12 years in the period 27 December 2020 - 30 September 2021 (the latest date for which outcome data were available). Regional health data were locally transformed into a study-specific Common Data Model and locally processed using *TheShinISS*.

In the end, regional pseudonymized datasets were provided to the National Institute of Health for centralized analysis, in compliance with EU General Data Protection Regulation. Over the last two years, *TheShinISS* framework has been employed in several large-scale observational studies exploring the association between some exposures and COVID-19 onset/prognosis as well as other drug and vaccine-related research topics and is currently maintained by a collaborative research network.^1,2,9-13^ The relational scheme of the study databases as well as TheShinISS flow diagram is described in Figure 1.

**Figure 1.**
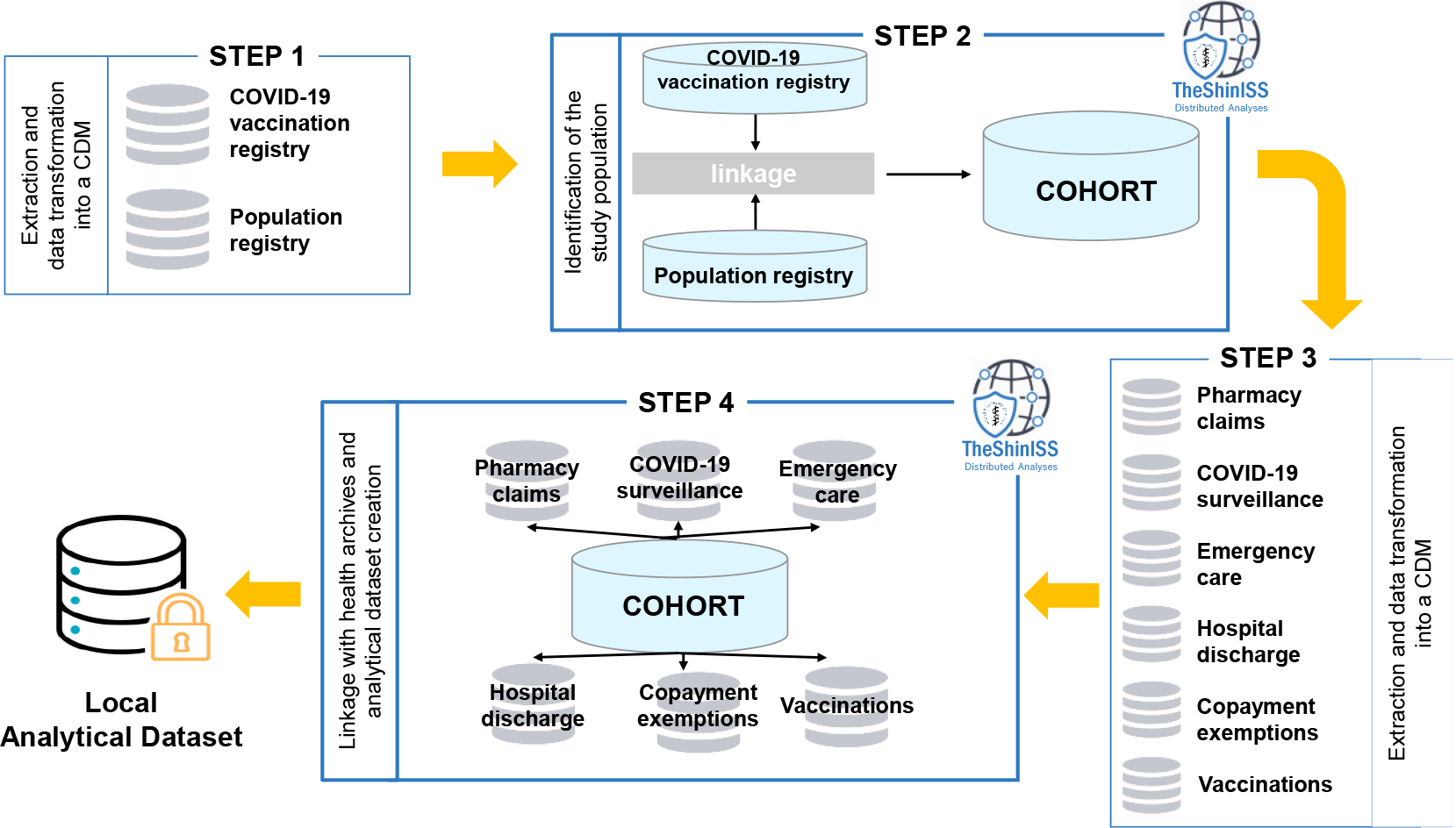
Diagram showing the data flow when using TheShinISS to locally process health care data structured according to a Common Data Model *CDM: Common Data Model*

Five Italian Regions (northern Italy: Lombardia, Veneto, Friuli Venezia Giulia and Emilia Romagna; central Italy: Lazio), representing 44% of the population aged ≥12 years resident in Italy, contributed data of all vaccinated persons in this age group, in a period ranging from 27/12/2020 to the last data update, which varied across Regions: Lombardia up to 30/09/2021, Veneto up to 20/06/2021, Friuli Venezia Giulia up to 31/08/2021, Emilia Romagna up to 30/06/2021 and Lazio up to 16/06/2021. We included in the study all persons aged ≥12 years who received at least a first dose of anti-COVID-19 vaccines and were admitted to emergency care or hospital with the outcome of GBS. We excluded individuals with missing or inconsistent information on relevant variables (age, sex, vaccine product and dose, date of vaccination, of death and of event). Furthermore, we excluded individuals with a history of GBS within 365 days leading up to the start of the study period.

The observation period for each case ranged from 27 December 2020 to the end of follow-up, which occurred at the end of Region-specific study period. If patients died, the end of the observation period was defined according to what is proposed by the SCCS methodology to handle mortality.^8^

### Definition of outcomes

The outcome of interest was the first diagnosis of GBS identified from emergency care and/or hospital admissions occurring during the observation period using International Classification of Disease, 9^th^ Revision, Clinical Modification (ICD-9-CM codes of GBS: 357.0).

### Definition of exposures

The exposures of interest were the first or second dose of BNT162b2 (Tozinameran), mRNA-1273 (Elasomeran), ChAdOx1-S and Ad26.COV2-S vaccines.

The exposure risk interval was defined as [0-42) days after first or second dose administration (vaccination date), which included day 0, the day of vaccination, according to Brighton Collaboration guidance.^14^ The unexposed baseline interval (reference period) was defined as any time of observation out of the risk intervals (before, between or after the risk intervals). According to the vaccination schedules of BNT162b2 (21-day interval between the first and second dose) and mRNA-1273 vaccines (28-day interval between the first and second dose), the 42-day risk intervals overlap and, consequently, the risk interval after first dose may end after the second dose. The convention in the SCCS methodology is that the most recent exposure period takes precedence over a previous exposure, and the parameterization of the SCCS model is adjusted accordingly.

### Statistical analysis

Characteristics of the cohort of vaccinated persons and GBS cases were described by age, sex, comorbidities and co-medications.

The SCCS model was fitted using unbiased estimating equations to estimate the Relative Incidences and their 95% Confidence Intervals (95% CI). In the following we will use the term “evidence of association” between vaccine exposure and the study event (overall and in a given subgroup) for a Relative Incidence estimate whose 95% CI does not include the null effect. To handle event-dependent exposures, the SCCS model was properly modified considering a counterfactual exposure history for any exposures arising after occurrence of an event.^3,7^ Six 45-day calendar periods were considered as time-varying covariate controlling for the seasonal effect. We also estimated the Excess of Cases (EC) per 100,000 vaccinated.^15^ We carried out subgroup analyses by age group (12-39, 40-59, ≥60 years), sex and vaccine product (BNT162b2, mRNA-1273, ChAdOx1-S and Ad26.COV2-S). We performed sensitivity analyses to assess the robustness of the results. We explored the seasonal effect by removing the calendar time factor, we investigated the effect of the SARS-CoV-2 infection by restricting the analyses to subjects without a positive SARS-CoV-2 test during the study period, and we explored the assumption that the most recent exposure period takes precedence over a previous exposure, fitting a common parameter for both doses. Moreover, to support the choice of the modified SCCS model, other sensitivity analyses were conducted using the standard SCCS method: beginning observation at time 0; beginning observation at exposure (starting the observation time at the first and second dose); including a [–28-0) day pre-risk period. The analyses were performed using R version 4.2.2 (R Core Team 2021) with SCCS package^16^ and STATA version 16.1.

### Ethics and permissions

This study was approved by the *National Unique Ethics Committee for the evaluation of clinical trials of medicines for human use and medical devices for patients with COVID-19 of the National Institute for Infectious Diseases “Lazzaro Spallanzan*i” in Rome (ordinance n. 335, 17/05/2021 and n. 399, 02/09/2021).

## Results

Among 27,889,821 doses uploaded on TheShinISS, the proportion of missing or inconsistent observations was 0.65% (n. 179,926) (**Figure 2**).

**Figure 2.**
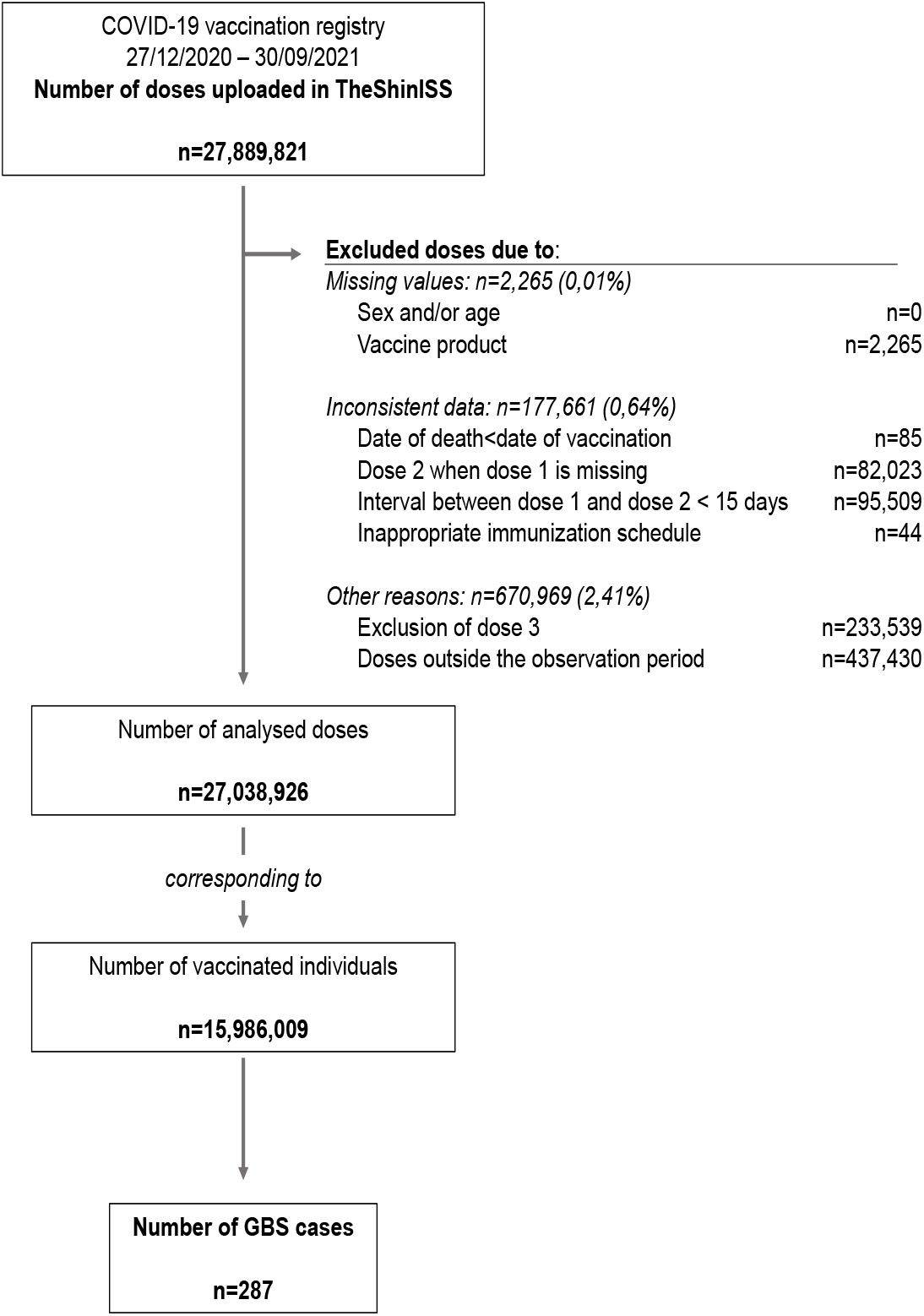
Flow chart of the study population

In the study period 27,038,926 first or second doses of anti-COVID-19 vaccines were administered to 15,986,009 persons aged 12 years or over in the 5 regions (**Figure 1, Table 1**), with a median follow-up time of 270 days (interquartile range 237-270 days). The overall vaccination coverage (VC) was 67.6% and exceeded 85% in those ≥60 years of age. The VC by region was: Lombardia (85.6%), Friuli Venezia Giulia (71.3%), Veneto (56.0%), Emilia Romagna (56.3%) and Lazio (54.3%). The highest VCs observed in Lombardia and Friuli Venezia Giulia depend on the vaccination registry update by region and therefore on the length of the observation period (month of last vaccination August-September vs June 2021 in the other regions).

**Figure 3.**
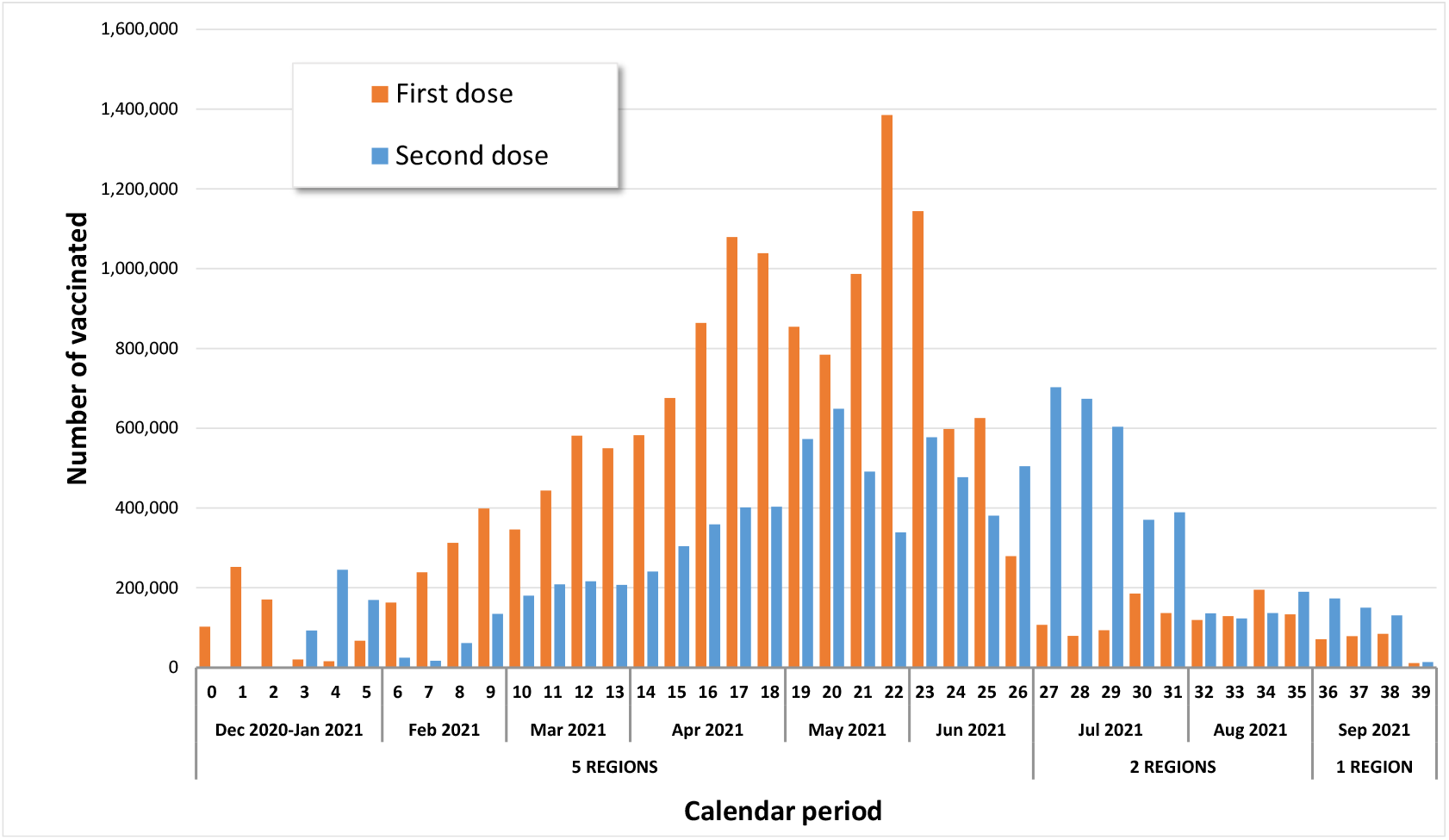
Distribution of first and second dose vaccine administrations by calendar week *Regional vaccination registry update: Veneto, Emilia Romagna, Lazio (Dec. 2020-Jun. 2021); Friuli Venezia Giulia (Dec. 2020-Aug. 2021); Lombardia (Dec. 2020-Sep. 2021)*

**Table 1.** Characteristics of vaccinated population by anti-COVID-19 vaccine brand (December 2020-September 2021)

| Vaccine brand | N. vaccinates | N. doses | Median age [IQ range] | M/F ratio |
| --- | --- | --- | --- | --- |
| BNT162b2 | 10,833,284 | 18,899,505 | 53 [40-69] | 0.91 |
| mRNA-1273 | 1,706,979 | 3,037,506 | 51 [36-69] | 0.94 |
| ChAdOx1-S | 2,863,950 | 4,520,119 | 64 [54-72] | 0.85 |
| Ad26.COVS-S | 581,796 | 581,796 | 57 [51-64] | 1.18 |
| **Number of subjects** | **15,986,009** | **27,038,926** | **56 [42-70]** | **0.91** |

*N.: number; IQ: interquartile; M/F: males/ females*

During the anti-COVID-19 vaccination campaign, in the 5 study regions, 4 different vaccines were used, two mRNA vaccines (BN162b2: n=10,833,284, 67.8%; mRNA-1273: n=1,706,979, 10.7%) and two viral vector vaccines (ChAdOx1-S: n=2,863,950, 17.9%; Ad26.COV2-S: n=581,796, 3.6%) (**Table 1**).

The 15,986,009 vaccinated subjects had a median age of 56 years, interquartile range (IQR) [42-70] and 52% were females. In **Table 2** are shown the clinical characteristics of the vaccinated subjects obtained from data available in the archives of pharmaceutical prescriptions (in the last 12 months), hospital admissions (in the last 5 years) and exemptions in the period prior to anti-COVID-19 vaccination.

**Table 2.**
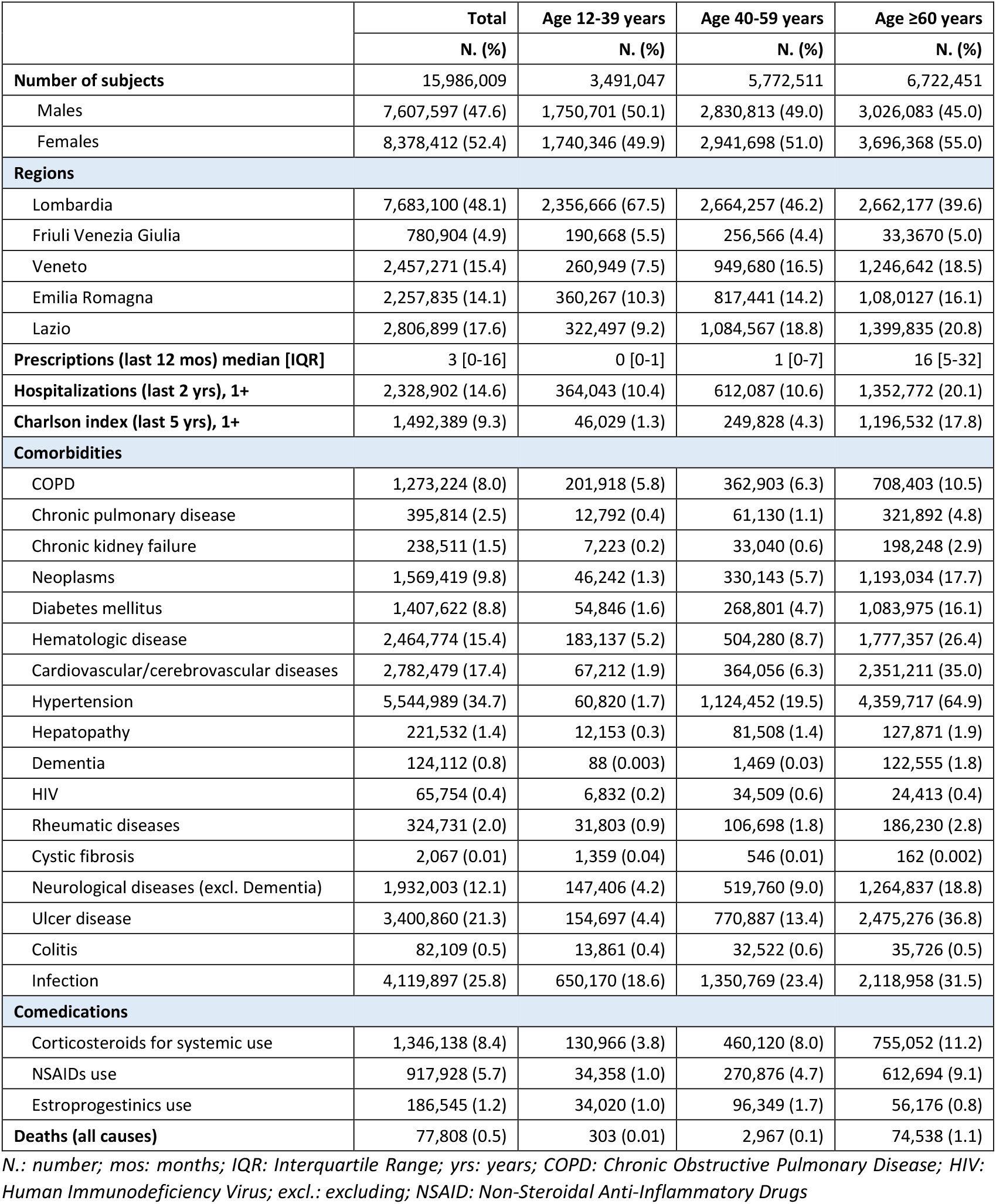
Characteristics of vaccinated population aged ≥12 years by age group (December 2020-September 2021)

| | Total | Age 12-39 years | Age 40-59 years | Age $\geq 60$ years |
| --- | --- | --- | --- | --- |
|  | N. (%) | N. (%) | N. (%) | N. (%) |
| **Number of subjects** | 15,986,009 | 3,491,047 | 5,772,511 | 6,722,451 |
| Males | 7,607,597 (47.6) | 1,750,701 (50.1) | 2,830,813 (49.0) | 3,026,083 (45.0) |
| Females | 8,378,412 (52.4) | 1,740,346 (49.9) | 2,941,698 (51.0) | 3,696,368 (55.0) |
| **Regions** |  |  |  |  |
| Lombardia | 7,683,100 (48.1) | 2,356,666 (67.5) | 2,664,257 (46.2) | 2,662,177 (39.6) |
| Friuli Venezia Giulia | 780,904 (4.9) | 190,668 (5.5) | 256,566 (4.4) | 33,3670 (5.0) |
| Veneto | 2,457,271 (15.4) | 260,949 (7.5) | 949,680 (16.5) | 1,246,642 (18.5) |
| Emilia Romagna | 2,257,835 (14.1) | 360,267 (10.3) | 817,441 (14.2) | 1,08,0127 (16.1) |
| Lazio | 2,806,899 (17.6) | 322,497 (9.2) | 1,084,567 (18.8) | 1,399,835 (20.8) |
| **Prescriptions (last 12 mos) median [IQR]** | 3 [0-16] | 0 [0-1] | 1 [0-7] | 16 [5-32] |
| **Hospitalizations (last 2 yrs), 1+** | 2,328,902 (14.6) | 364,043 (10.4) | 612,087 (10.6) | 1,352,772 (20.1) |
| **Charlson index (last 5 yrs), 1+** | 1,492,389 (9.3) | 46,029 (1.3) | 249,828 (4.3) | 1,196,532 (17.8) |
| **Comorbidities** |  |  |  |  |
| COPD | 1,273,224 (8.0) | 201,918 (5.8) | 362,903 (6.3) | 708,403 (10.5) |
| Chronic pulmonary disease | 395,814 (2.5) | 12,792 (0.4) | 61,130 (1.1) | 321,892 (4.8) |
| Chronic kidney failure | 238,511 (1.5) | 7,223 (0.2) | 33,040 (0.6) | 198,248 (2.9) |
| Neoplasms | 1,569,419 (9.8) | 46,242 (1.3) | 330,143 (5.7) | 1,193,034 (17.7) |
| Diabetes mellitus | 1,407,622 (8.8) | 54,846 (1.6) | 268,801 (4.7) | 1,083,975 (16.1) |
| Hematologic disease | 2,464,774 (15.4) | 183,137 (5.2) | 504,280 (8.7) | 1,777,357 (26.4) |
| Cardiovascular/cerebrovascular diseases | 2,782,479 (17.4) | 67,212 (1.9) | 364,056 (6.3) | 2,351,211 (35.0) |
| Hypertension | 5,544,989 (34.7) | 60,820 (1.7) | 1,124,452 (19.5) | 4,359,717 (64.9) |
| Hepatopathy | 221,532 (1.4) | 12,153 (0.3) | 81,508 (1.4) | 127,871 (1.9) |
| Dementia | 124,112 (0.8) | 88 (0.003) | 1,469 (0.03) | 122,555 (1.8) |
| HIV | 65,754 (0.4) | 6,832 (0.2) | 34,509 (0.6) | 24,413 (0.4) |
| Rheumatic diseases | 324,731 (2.0) | 31,803 (0.9) | 106,698 (1.8) | 186,230 (2.8) |
| Cystic fibrosis | 2,067 (0.01) | 1,359 (0.04) | 546 (0.01) | 162 (0.002) |
| Neurological diseases (excl. Dementia) | 1,932,003 (12.1) | 147,406 (4.2) | 519,760 (9.0) | 1,264,837 (18.8) |
| Ulcer disease | 3,400,860 (21.3) | 154,697 (4.4) | 770,887 (13.4) | 2,475,276 (36.8) |
| Colitis | 82,109 (0.5) | 13,861 (0.4) | 32,522 (0.6) | 35,726 (0.5) |
| Infection | 4,119,897 (25.8) | 650,170 (18.6) | 1,350,769 (23.4) | 2,118,958 (31.5) |
| **Comedications** |  |  |  |  |
| Corticosteroids for systemic use | 1,346,138 (8.4) | 130,966 (3.8) | 460,120 (8.0) | 755,052 (11.2) |
| NSAIDs use | 917,928 (5.7) | 34,358 (1.0) | 270,876 (4.7) | 612,694 (9.1) |
| Estroprogestinics use | 186,545 (1.2) | 34,020 (1.0) | 96,349 (1.7) | 56,176 (0.8) |
| **Deaths (all causes)** | 77,808 (0.5) | 303 (0.01) | 2,967 (0.1) | 74,538 (1.1) |

N.: number; mos: months; IQR: Interquartile Range; yrs: years; COPD: Chronic Obstructive Pulmonary Disease; HIV: Human Immunodeficiency Virus; excl.: excluding; NSAID: Non-Steroidal Anti-Inflammatory Drugs

In **Table 3** are shown the characteristics of the GBS cases. Over the observation period, among 15,986,009 vaccinees, 287 had a new diagnosis of GBS (median age 65 years, IQR [54-76]), 184 cases (64.1%) after the first dose of anti-COVID-19 vaccine. Among 10,833,284 recipients of BNT162b2 vaccine, 187 had GBS. Among 1,706,979 recipients of mRNA-1273, 25 individuals had GBS. Among 2,863,950 ChAdOx1-S vaccinees and 581,796 Ad26.COV2-S vaccinees there were 58 and 17 new cases of GBS. Thirteen deaths for all causes of death were observed during the study observation period, with a median age of 80 years, IQR [74-81] and 54% with a Charlson Index greater than 1.

**Table 3.** Characteristics of GBS cases by vaccine brand

|  | N. of events (%) |  |  |  |  |
| --- | --- | --- | --- | --- | --- |
|  | Total | BNT162b2 | mRNA-1273 | ChAdOx1-S | Ad26.COVS-S |
| **Number of subjects** | **287** | **187** | **25** | **58** | **17** |
| **Sex** |  |  |  |  |  |
| Males | 171 (59.6) | 113 (60.4) | 16 (64.0) | 32 (55.2) | 10 (58.8) |
| Females | 116 (40.4) | 74 (39.6) | 9 (36.0) | 26 (44.8) | 7 (41.2) |
| **Age (years)** |  |  |  |  |  |
| median [IQR] | 65 [54-76] | 65 [52-78] | 64 [54-78] | 68 [59-72] | 63 [55-68] |
| <40 | 26 (9.1) | 25 (13.4) | - | - | 1 (5.9) |
| 40-59 | 76 (26.5) | 47 (25.1) | 10 (40.0) | 15 (25.9) | 4 (23.5) |
| ≥60 | 185 (64.5) | 115 (61.5) | 15 (60.0) | 43 (74.1) | 12 (70.6) |
| **Regions** |  |  |  |  |  |
| Lombardia | 144 (50.2) | 99 (52.9) | 13 (52.0) | 26 (44.8) | 6 (35.3) |
| Friuli Venezia Giulia | 20 (7.0) | 10 (5.3) | 3 (12.0) | 6 (10.3) | 1 (5.9) |
| Veneto | 31 (10.8) | 22 (11.8) | 1 (4.0) | 6 (10.3) | 2 (11.8) |
| Emilia Romagna | 50 (17.4) | 30 (16.0) | 7 (28.0) | 7 (12.1) | 6 (35.3) |
| Lazio | 42 (14.6) | 26 (13.9) | 1 (4.0) | 13 (22.4) | 2 (11.8) |
| **Deaths** | **13 (4.5)** | **10 (5.3)** | **2 (8.0)** | **1 (1.7)** | **-** |
| **N. events ≥ first dose** | **184 (64.1)** | **103 (55.1)** | **15 (60.0)** | **55 (94.8)** | **11 (64.7)** |
| **N. positive SARS-CoV-2 test** | **33 (11.5)** | **21 (11.2)** | **5 (20.0)** | **3 (5.2)** | **4 (23.5)** |

**Table 3 cont.** Characteristics of GBS cases by vaccine brand
|  | N. of events (%) |  |  |  |  |
| --- | --- | --- | --- | --- | --- |
|  | Total | BNT162b2 | mRNA-1273 | ChAdOx1-S | Ad26.COVS-2 |
| **Number of subjects** | **287** | **187** | **25** | **58** | **17** |
| Prescriptions (last 12 mos), median [IQR] | 12 [3-28] | 15 [5-31] | 16 [5-39] | 8 [1-23] | 3 [0-6] |
| Hospitalizations (last 2 yrs), 1+ | 135 (47.0) | 112 (59.9) | 12 (48.0) | 7 (12.1) | 4 (23.5) |
| Charlson index (last 5 yrs), 1+ | 134 (46.7) | 95 (50.8) | 18 (72.0) | 18 (31.0) | 3 (17.6) |
| **Comorbidities** |  |  |  |  |  |
| COPD | 30 (10.5) | 17 (9.1) | 8 (32.0) | 5 (8.6) | - |
| Chronic pulmonary disease | 49 (17.1) | 37 (19.8) | 7 (28.0) | 2 (3.4) | 3 (17.6) |
| Chronic kidney failure | 10 (3.5) | 7 (3.7) | 3 (12.0) | - | - |
| Neoplasms | 50 (17.4) | 37 (19.8) | 5 (20.0) | 6 (10.3) | 2 (11.8) |
| Diabetes mellitus | 50 (17.4) | 36 (19.3) | 6 (24.0) | 8 (13.8) | - |
| Hematologic disease | 86 (30.0) | 67 (35.8) | 9 (36.0) | 8 (13.8) | 2 (11.8) |
| Cardiovascular/cerebrovascular diseases | 95 (33.1) | 73 (39.0) | 9 (36.0) | 10 (17.2) | 3 (17.6) |
| Hypertension | 163 (56.8) | 110 (58.8) | 17 (68.0) | 30 (51.7) | 6 (35.3) |
| Hepatopathy | 6 (2.1) | 2 (1.1) | 4 (16.0) | - | - |
| Dementia | 2 (0.7) | 2 (1.1) | - | - | - |
| HIV | 1 (0.3) | - | 1 (4.0) | - | - |
| Rheumatic diseases | 7 (2.4) | 6 (3.2) | 1 (4.0) | - | - |
| Cystic fibrosis | - | - | - | - | - |
| Neurological diseases (excl. Dementia) | 80 (27.9) | 60 (32.1) | 12 (48.0) | 8 (13.8) | - |
| Ulcer disease | 108 (37.6) | 87 (46.5) | 8 (32.0) | 11 (19.0) | 2 (11.8) |
| Colitis | 3 (1.0) | 3 (1.6) | - | - | - |
| Infection | 103 (35.9) | 77 (41.2) | 8 (32.0) | 15 (25.9) | 3 (17.6) |
| **Comedications** |  |  |  |  |  |
| Corticosteroids for systemic use | 65 (22.6) | 47 (25.1) | 7 (28.0) | 6 (10.3) | 5 (29.4) |
| NSAIDs use | 29 (10.1) | 21 (11.2) | 1 (4.0) | 5 (8.6) | 2 (11.8) |
| Estroprogestinics use | 2 (0.7) | 2 (1.1) | - | - | - |

N.: number; mos: months; IQR: Interquartile Range; yrs: years; COPD: Chronic Obstructive Pulmonary Disease; HIV: Human Immunodeficiency Virus; excl.: excluding; NSAID: Non-Steroidal Anti-Inflammatory Drugs

During the 42-day risk interval there were a total of 67 cases of GBS after the first dose and 41 cases after the second dose (**Table 4**). In the 42-day risk interval, increased risks were observed after the administration of first dose (RI=6.83; 95% CI 2.14-21.85) and second dose (RI=7.41; 95% CI 2.35-23.38) for mRNA-1273 vaccine, corresponding to estimated 0.4 and 0.3 EC per 100,000 vaccinated respectively. Considering the overlapping of the 42-day risk periods between the first and second dose for mRNA-1273 vaccine, it cannot be excluded that the increased risk observed in the 42-day risk period after the second dose might be partially driven by the effect of the first dose. Increased risk was also observed after the first dose of ChAdOx1-S vaccine (RI=6.52; 95% CI 2.88-14.77), corresponding to estimated 1.0 EC per 100,000 vaccinated. There was no evidence of increased risk of GBS after vaccination with BNT162b2 and Ad26.COV2-S vaccines (**Table 4**).

**Table 4.** Relative Incidences estimated by Self-Controlled Case Series model by vaccine brand and dose: 287 Guillain Barré Syndrome events in the anti-COVID-19 vaccinated population

|  | Dose | Risk period | BNT162b |  | mRNA-1273<sup>^</sup> |  | ChAdOx1-S |  | Ad26.COV2-S |  |
| --- | --- | --- | --- | --- | --- | --- | --- | --- | --- | --- |
|  |  |  | Events (n. 187) | RI (95% CI) | Events (n. 25) | RI (95% CI) | Events (n. 58) | RI (95% CI) | Events (n. 17) | RI (95% CI) |
| **Total** |  | *Ref.* | 138 | 1 | 13 | 1 | 18 | 1 | 10 | 1 |
|  | 1 | [0-42) | 19 | 0.85 (0.49-1.48) | 7 | 6.83 (2.14-21.85) | 34 | 6.52 (2.88-14.77) | 7 | 1.94 (0.32-11.69) |
|  | 2 | [0-42) | 30 | 1.30 (0.80-2.10) | 5 | 7.41 (2.35-23.38) | 6 | 3.56 (0.31-40.29) |  |  |
| **Males** |  | *Ref.* | 89 | 1 | 8 | 1 | 11 | 1 | 6 | 1 |
|  | 1 | [0-42) | 9 | 0.64 (0.30-1.36) | 3 | 5.26 (0.94-29.42) | 16 | 4.94 (1.84-13.28) | 4 | 1.06 (0.15-7.39) |
|  | 2 | [0-42) | 15 | 1.06 (0.56-2.00) | 5 | 16.50 (3.01-90.56) | 5 | 1.54 (0.13-18.39) |  |  |
| **Females** |  | *Ref.* | 49 | 1 | 5 | 1 | 7 | 1 | 4 | 1 |
|  | 1 | [0-42) | 10 | 1.19 (0.55-2.55) | 4 | 13.44 (2.83-63.80) | 18 | 7.14 (1.94-26.19) | 3 | 2.35 (0.18-30.00) |
|  | 2 | [0-42) | 15 | 1.84 (0.85-4.00) | 0 | - | 1 | * |  |  |
| **12-39 yrs** |  | *Ref.* | 18 | 1 | 0 | 1 | 0 | 1 | 1 | 1 |
|  | 1 | [0-42) | 4 | 0.68 (0.15-3.14) | 0 | - | 0 | - | 0 | - |
|  | 2 | [0-42) | 3 | 0.64 (0.13-3.29) | 0 | - | 0 | - |  |  |
| **40-59 yrs** |  | *Ref.* | 34 | 1 | 6 | 1 | 4 | 1 | 2 | 1 |
|  | 1 | [0-42) | 6 | 1.02 (0.36-2.84) | 3 | * | 10 | 4.50 (1.37-14.79) | 2 | * |
|  | 2 | [0-42) | 7 | 1.63 (0.61-4.37) | 1 | * | 1 | * |  |  |
| **≥60 yrs** |  | *Ref.* | 86 | 1 | 7 | 1 | 14 | 1 | 7 | 1 |
|  | 1 | [0-42) | 9 | 0.74 (0.35-1.57) | 4 | 8.03 (2.08-31.03) | 24 | 6.84 (2.56-18.28) | 5 | 1.78 (0.27-11.58) |
|  | 2 | [0-42) | 20 | 1.21 (0.65-2.25) | 4 | 7.71 (2.38-24.97) | 5 | 1.97 (0.13-30.39) |  |  |

RI: Relative Incidence; CI: Confidence Interval; n.: number; Ref.: reference; yrs: years
<sup>^</sup>3 calendar periods of 90 days; \* RI not estimable by the SCCS model

### Subgroup and sensitivity analyses

In the subgroup analysis by sex (**Table 4**), an increased risk of GBS was observed in the 0-42 days risk period among both males and females after mRNA-1273 vaccine. More specifically, in males an increased risk was observed after the first dose, with a borderline significance (RI=5.26; 95% CI 0.94-29.42; p=0.06) and the second dose (RI=16.50; 95% CI 3.01-90.56); in females after the first dose (RI=13.44; 95% CI 2.83-63.80). There was also evidence of an increased risk after a first dose of ChAdOx1-S in males (RI=4.94; 95% CI 1.84-13.28) and females (RI=7.14; 95% CI 1.94-26.19).

In the subgroup analysis by age (**Table 4**), there was evidence of an increased risk of GBS with mRNA-1273 vaccine among those aged ≥60 years after the first (RI=8.03; 95% CI 2.08-31.03) and second dose (RI=7.71; 95% CI 2.38-24.97). After a first dose of ChAdOx1-S there was evidence of an increased risk of GBS in those aged 40-59 (RI=4.50; 95% CI 1.37-14.79) and in those aged ≥60 years (RI=6.84; 95% CI 2.56-18.28). There was no evidence of increased risk of GBS after vaccination with BNT162b2 and Ad26.COV2-S vaccines in the subgroup analysis by age and sex.

Due to the low number of cases, the analyses in the sub-divided risk-periods ([0-14), [14-28), [28-42) days) were limited to BNT162b vaccine. This analysis confirms the lack of association seen in the 42-day risk period.

All sensitivity analyses, performed to assess the robustness of the SCCS methodology, were consistent with the main results (**Table 5**). More specifically, it is reassuring that the sensitivity analysis starting the observation time at the exposure, in the standard SCCS method, gave essentially similar point estimates to our main analysis.

**Table 5.**
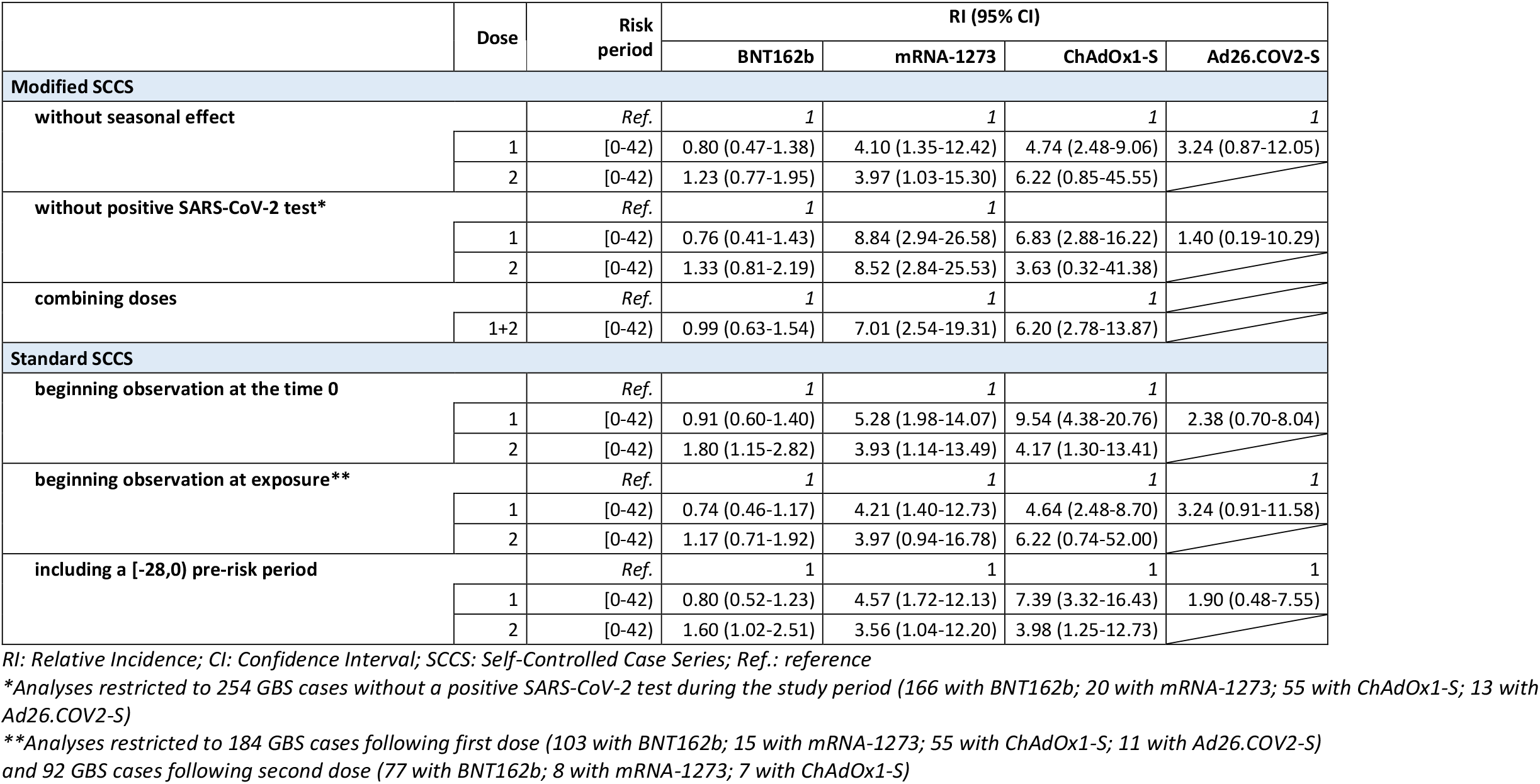
Sensitivity analyses: Relative Incidences estimated by Self-Controlled Case Series model by vaccine brand and dose (287 Guillain Barré Syndrome events in the anti-COVID-19 vaccinated population)

## Discussion

This SCCS study, covering about 16 million people, found evidence of an increased risk of GBS after administration of first and second dose of mRNA-1273 and first dose of ChAdOx1-S in the risk period of 0-42 days. However, we did not observe an increased risk of GBS following first and second dose of BNT162b nor any increased risk after Ad26.COV2-S vaccination. Assuming a causal effect, the number of the estimated EC were low with 0.4 and 0.3 EC per 100,000 vaccinated for first and second dose of mRNA-1273 and 1.0 EC per 100,000 vaccinated for the first dose of ChAdOx1-S.

Subgroup analyses by sex found an increased risk of GBS with mRNA-1273 after the second dose in males (with a borderline statistical significance after the first dose) and after the first dose in females. An increased risk was also observed after the first dose of ChAdOx1-S both in males and females. Stratifying data by age, an increased risk was observed in those aged 40-59 years after first dose of ChAdOx1-S. Additionally, we found evidence of an association with GBS of both mRNA-1273 and ChAdOx1-S vaccines in those aged ≥60 years.

Our results are in lines with previous findings from surveillance studies in England with SCCS design^17,18^ investigating the association between ChAdOx1-S and BNT162b vaccines with acute neurological outcomes. For example, Walker *et al*.^17^ found an increased incidence of GBS after a first dose of ChAdOx1-S at 4-42 days post vaccination (Incidence Rate Ratio=2.85; 95% CI 2.33-3.47) and no evidence of an association after first and second dose for BNT162b. This study did not investigate the relationship between mRNA-1273 and GBS due to limited power. Similarly, the study of Patone *et al*.^18^ observed an increased risk of GBS following first dose of ChAdOx1-S at 1-28 days after vaccination (Incidence Rate Ratio=2.04; 95% CI 1.60-2.60) but nor after first dose of BNT162b vaccination. This study had insufficient follow-up to assess the effect of the second dose of COVID-19 vaccines.

Our study has the strengths of combining larger study population and longer follow-up than previous studies which permitted us to obtain unbiased estimates with the SCCS model for the first and second dose of mRNA-1273. Further strengths are the broad geographical distribution of the cohort as well as the availability of accurate and exhaustive data on COVID-19 vaccination and outcome. Another strength is the robustness of the SCCS method applied which is not susceptible to confounding by known and unknown factors that are time-invariant during the study period. The seasonality effect was also included in the models as an important time variant confounding factor. We also conducted a number of sensitivities analyses to investigate the robustness of our results to the assumptions of SCCS model.

## Conclusions

With the implementation of a worldwide COVID-19 vaccination campaign it is important the continuous monitoring of the suspected adverse events of these new vaccines as key component of any vaccination program. The aim of the pharmacovigilance activities is to provide evidence for benefit-risk profile of vaccination and, also importantly, to provide reassurance about the rarity of the events.

Our population-based study, found an increased risk of GBS after administration of first and second dose of mRNA-1273 and first dose of ChAdOx1-S. However, these findings were compatible with a small number of excess cases. Age-specific estimates indicated an increased risk in those aged 40-59 years and aged ≥60 years vaccinated with ChAdOx1-S. Additionally, an increased risk of GBS was found with mRNA-1273 in those aged ≥60 years.

Our data are reassuring regarding BNT162b and Ad26.COV2-S vaccines with respect to GBS outcome. No increased risk of GBS was detected following each of BNT162b vaccine dose nor any increased risk after Ad26.COV2-S vaccine dose. These findings are likely to be of relevance to regulators, health professionals and developers of clinical guidelines in the risk benefit evaluations of the COVID-19 vaccines.

## Data Availability

Data cannot be shared publicly under article 9 of Regulation (EU) 2016/679. Data are available from the Data Protection Officer of Istituto Superiore di Sanità Dott. Carlo Villanacci, for researchers who meet the criteria for access to confidential data.

## Data availability statement

Data cannot be shared publicly under article 9 of Regulation (EU) 2016/679. Data are available from the Data Protection Officer of Istituto Superiore di Sanità-Dott. Carlo Villanacci, for researchers who meet the criteria for access to confidential data.

## Acknowledgements

We would like to thank Gianpaolo Scalia Tomba (University of Tor Vergata, Rome) for methodological advice. We would like to thank members of TheShinISS-vax|COVID Surveillance Group collaborating to this study: Maria Cutillo, Ilaria Ippoliti, Giuseppe Marano, Flavia Mayer (National Centre for Drug Research and Evaluation, National Institute of Health - Istituto Superiore di Sanità); Michele Ercolanoni (Lombardia Region); Ugo Moretti, Giovanna Scroccaro, Paola Deambrosis, Manuel Zorzi, Sara Contin, Michele Tonon, Elena Vecchiato (Veneto Region); Paola Rossi, Sara Samez (Friuli Venezia Giulia Region); Nazanin Morgheiseh (Emilia Romagna Region); Lorella Lombardozzi, Valeria Desiderio, Maria Balducci, Francesca Romana Poggi (Lazio Region).

## Footnotes

### Contributors

CM and SSA contributed equally as joint first authors. CM, SSA, FMI, VB, GT, RDC and MM conceived the study question and designed the study. CM, SSA, VB, AP, EC, EB and MM retrieved and prepared the data. CM, SSA and MM carried out the analysis. CM, SSA, FMI, VB, GT, GZ, ES, NM, OL, GM, CZ, RDC and MM interpreted data. CM, SSA and MM wrote the first draft of the manuscript. CM, SSA, FMI, VB, GT, GZ, AP, ES, NM, OL, GM, EC, CZ, EB, RDC and MM critically revised the paper. All authors contributed to subsequent drafts and interpretation of the findings and approved the final version of the manuscript. CM, SSA and MM are manuscript’s guarantors. The corresponding author (MM) attests that all listed authors meet authorship criteria and that no others meeting the criteria have been omitted.

## Funding

The Istituto Superiore di Sanità received funding from AIFA (Italian Medicines Agency) www.aifa.gov.it for this study in the framework of the collaboration agreement “Efficacia real world e sicurezza dei vaccini anti Covid-19: studio di coorte e Self-Controlled Case Series” (Effectiveness and safety of COVID-19 vaccines: cohort and Self-Controlled Case Series studies). AIFA is the Italian national regulatory body for drugs and vaccines and a public organization. All authors are independent from the funder. The funders had no role in study design, data collection and analysis, decision to publish, or preparation of the manuscript.

## Competing interests

All authors have completed the ICMJE uniform disclosure form at www.icmje.org/disclosureof-interest/. CM, SSA, FMI, AP, ES, VB, NM, OL, EC, CZ, RDC and MM have no conflicts of interest to disclose. GZ, GM report participation in the last 36 months on Working Group on Safety signals analysis at the Agenzia Italiana del Farmaco (Italian Medicine Agency).

GT coordinated a pharmacoepi team at the University of Messina till Oct 2020 and currently at the academic spin-off INSPIRE that received research grants from PTC Therapeutics, Kiowa Kirin, Chiesi, Daiichi Sankyo for the conduct of observational studies on topics not related to the paper; GT participated to Advisory Board/interview sponsored by Eli Lilly, Amgen, Sanofi, SOBI, Gilead, ABBvie, Verpora and Daiichi Sankyo on topics not related to the paper.

## Notes

### Competing Interest Statement

The authors have declared no competing interest.

### Author Declarations

This study was approved by the National Unique Ethics Committee for the evaluation of clinical trials of medicines for human use and medical devices for patients with COVID 19 of the National Institute for Infectious Diseases Lazzaro Spallanzani in Rome (ordinance n. 335, 17/05/2021 and n. 399, 02/09/2021).

### Summary of Updates

Correction of the Title to appear properly the accent of the word Barre Inclusion of one missed author (Emanuela Bovo) and one member of TheShinISS vax COVID Surveillance Group in the Acknowledgements section (Sara Contin)

## References

1. Massari M, Spila Alegiani S, Morciano C, Spuri M, Marchione P, Felicetti P, et al. Post-marketing active surveillance of myocarditis and pericarditis following vaccination with COVID-19 mRNA vaccines in persons aged 12-39 years in Italy: a multi-database, self-controlled case series study. PLoS Med 2022; 19(7): e1004056. https://doi.org/10.1371/journal.pmed.1004056

2. Massari M, Spila Alegiani S, Da Cas R, Menniti Ippolito F. TheShinISS: an open-source tool for conducting distributed analyses within pharmacoepidemiological multi-database studies. Boll Epidemiol Naz 2020;1(2):39–45. doi:10.53225/BEN_006

3. Whitaker HJ, Farrington CP, Spiessens B, Musonda P. Tutorial in bio-statistics: the self-controlled case series method. Stat Med 2006;25(10):1768–97. doi:10.1002/sim.2302

4. Petersen I, Douglas I, Whitaker H. Self controlled case series methods: an alternative to standard epidemiological study designs. BMJ 2016;354:i4515. doi:10.1136/bmj.i4515.

5. Weldeselassie YG, Whitaker HJ, Farrington CP. Use of the self-controlled case-series method in vaccine safety studies: review and recommendations for best practice. Epidemiol Infect 2011;139 (12):1805–17. doi:10.1017/S0950268811001531

6. Farrington CP, Whitaker HJ, Hocine MN. Case series analysis for censored, perturbed, or curtailed post-event exposures. Biostatistics 2009;10(1):3–16. doi:10.1093/biostatistics/kxn013

7. Farrington CP, Whitaker H, Weldeselassie YG. Self-Controlled Case Series Studies. A Modelling Guide with R. CRC Press, 2018

8. Ghebremichael-Weldeselassie Y, Jabagi MJ, Botton J, Bertrand M, Baricault B, Drouin J, et al. A modified self-controlled case series method for event-dependent exposures and high event-related mortality, with application to COVID-19 vaccine safety. [published online ahead of print, 2022 Jan 28]. Stat Med 2022;10.1002/sim.9325. doi:10.1002/sim.9325

9. Trifirò G, Massari M, Da Cas R, Menniti Ippolito F, Sultana J, Crisafulli S, et al. RAAS inhibitor group. Renin–Angiotensin–Aldosterone system inhibitors and risk of death in patients hospitalised with covid19: a retrospective Italian cohort study of 43,000 patients. Drug Saf 2020; 43(12):1297–1308. doi:10.1007/s40264-020-00994-5

10. Spila Alegiani S, Crisafulli S, Giorgi Rossi P, Mancuso P, Salvarani C, Atzeni F, et al. Risk of COVID-19 hospitalization and mortality in rheumatic patients treated with hydroxychloroquine or other conventional DMARDs in Italy. Rheumatology (Oxford). 2021 Oct 9;60(SI):SI25–SI36. doi: 10.1093/rheumatology/keab348. PMID: 33856453; PMCID: PMC8083276

11. Massari M, Spila Alegiani S, Fabiani M, Belleudi V, Trifirò G, Kirchmayer U, et al. Association of influenza vaccination and prognosis in patients testing positive to SARS-COV-2 swab test: a largescale Italian multi-database cohort study. Vaccines 2021, 9(7), 716; https://doi.org/10.3390/vaccines9070716

12. Trifirò G, Isgrò V, Ingrasciotta Y, Ientile V, L’Abbate L, Foti SS, et al. Large-scale postmarketing surveillance of biological drugs for immune-mediated inflammatory diseases through an Italian distributed multi-database healthcare network: the VALORE Project. BioDrugs 2021;35(6):749–764. doi:10.1007/s40259-021-00498-3.

13. Belleudi V, Rosa AC, Finocchietti M, Poggi FR, Marino ML, Massari M, et al. An Italian multicentre distributed data research network to study the use, effectiveness, and safety of immunosuppressive drugs in transplant patients: Framework and perspectives of the CESIT project. Front. Pharmacol. 2022 13:959267. doi: 10.3389/fphar.2022.959267

14. AESI Case Definition Companion Guide for 1st Tier AESI. Guillain Barré and Miller Fisher Syndromes. Work Package: WP2 Standards and tools. V1.0 – February 9th, 2021. Authors: Barbara Law. Nature: Report | Diss. level: Public. https://brightoncollaboration.us/wp-content/uploads/2021/03/SPEAC_D2.5.2.1-GBS-Case-Definition-Companion-Guide_V1.0_format12062-1.pdf

15. Wilson K, Hawken S. Drug safety studies and measures of effect using the self-controlled case series design. Pharmacoepidemiol Drug Saf 2013;22:108–110. doi:10.1002/pds.3337

16. Weldeselassie YJ, Whitaker H and Farrington P (2021). SCCS: The Self-Controlled Case Series Method. R. package version 1.5. Available from: https://CRAN.R-project.org/package=SCCS

17. Walker JL, Schultze A, Tazare J, Tamborska A S. Safety of COVID-19 vaccination and acute neurological events: A self-controlled case series in England using the OpenSAFELY platform. Vaccine. 2022 Jul 30;40(32):4479–4487 doi: 10.1016/j.vaccine.2022.06.010.

18. Patone M, Handunnetthi L, Saatci D, Pan J, Katikireddi SV, Razvi S, Hunt D, Mei XW, Dixon S, Zaccardi F, Khunti K, Watkinson P, Coupland CAC, Doidge J, Harrison DA, Ravanan R, Sheikh A, Robertson C, Hippisley-Cox J. Neurological complications after first dose of COVID-19 vaccines and SARS-CoV-2 infection. Nat Med. 2021 Dec;27(12):2144–2153. doi: 10.1038/s41591-021-01556-7.

